# Minimising exposure to droplet and aerosolised pathogens: a computational fluid dynamics study

**DOI:** 10.1101/2020.05.30.20117671

**Authors:** Paolo Perella, Mohammad Tabarra, Ertan Hataysal, Amir Pournasr, Ian Renfrew

## Abstract

**Background:** Hazardous pathogens are spread in either droplets or aerosols produced during aerosol generating procedures (AGP). Adjuncts minimising exposure of healthcare workers to hazardous pathogens released during AGP may be beneficial. We used state-of-the-art Computational Fluid Dynamics modelling to optimise the performance of a custom-designed shield.

**Methods:** We modelled airflow patterns and trajectories of particles (size range 1–500µm) emitted during a typical cough using Computational Fluid Dynamics (ANSYS Fluent software), in the presence and absence of a protective shield enclosing the head of a patient. We modelled the effect of different shield designs, suction tube position, and suction flow rate on particle escape from the shield.

**Results:** Use of the shield prevented escape of 99.1–100% of particles, which were either trapped on the shield walls (16–21%) or extracted via suction (79–82%). At most, 0.9% particles remained floating inside the shield. Suction flow rates (40–160L min^−1^) had no effect on the final location of particles in a closed system. Particle removal from within the shield was optimal when a suction catheter was placed vertically next to the head of the patient. Addition of multiple openings in the shield reduced the purging performance from 99% at 160 L min^−1^ to 67% at 40 L min^−1^.

**Conclusion:** Computational fluid dynamics modelling provides information to guide optimisation of the efficient removal of hazardous pathogens released during AGP from a custom-designed shield. These data are essential to establish before clinical use and/or pragmatic clinical trials.

## Introduction

Hazardous respiratory pathogens are transmitted in droplet form, aerosols or by fomite deposition on surfaces^1^. Minimising these transmission routes is likely to reduce the exposure of healthcare workers to dangerous pathogens, including COVID-19. Coughing releases droplets and aerosolised particles with varied diameters and speeds^2^ which settle on local surfaces (droplets) or remain aerosolised. Understanding the physics of droplets and airflow can aid the design and guide use of equipment and systems to protect staff and populations against transmissible disease. Computational Fluid Dynamics (CFD) is a specialist area of mathematics and fluid mechanics used to solve complex engineering problems in industries including aerospace, aviation and construction. CFD has been used to model cardiorespiratory therapies (e.g. right ventricular assist devices^3^ and the design of operating rooms^4^). The predictive success of CFD modelling has obviated the reliance on wind tunnel testing in aviation and racing car design. CFD has also been used successfully in modelling biocontamination. The performance of CFD in predicting simulated aerosolised microbial (Bacillus licheniformis/aerius) deposition matched the final location of actual microbes on the internal surfaces of a spacecraft^5^.

To date, the effect of using shields to minimise aerosol contamination in AGP has not been rigorously tested. We used CFD modelling to test and optimise the performance of a protective shield designed to minimize the spread of droplets and smaller particles produced during AGP, an essential prerequisite before clinical use and/or the assessment of such devices in pragmatic clinical trials.

## Methods

### Shield design

In collaboration with Rolls-Royce and the Manufacturing Technology Centre (MTC), we designed a shield to reduce both Healthcare Worker (HCW) exposure and local environmental contamination (Figure 1). The shield is lightweight (4 kg) and easy to manoeuvre. Access ports on each side and vertical access ports are covered by overlapping silicone which forms a seal during use (Figure 1). The silicone flaps are soft to avoid damaging PPE worn by the operators. A silicone curtain lies over the entrance to the shield, containing both droplets and aerosols within the shield. At the base, 5cm ports on each side enable suction and ventilation tubing to be maintained at all times and closed loop ventilation to continue once shield is removed. Further details and video can be found in the supplementary material. When empty, the shield has a volume of 142.8 litres, which reduces to 108 litres once the shield is placed over a patient. With an extraction rate of 40 L min^−1^, 22 air changes per hour (ach) are achieved, rising to 89ach at an extraction rate of 160 L min^−1^. Given such large air change inside the box relative to the room, ambient air is drawn into the box. Experiments 1–3 were modelled on the assumption that ambient pressure around the box was 1 atm. In experiment 4, a room of dimensions 4m x 4.5m x 3.1m, with air change rate of 10ach and an ambient temperature of 23°C, was modelled (see supplementary material for further details).

**Figure 1.**
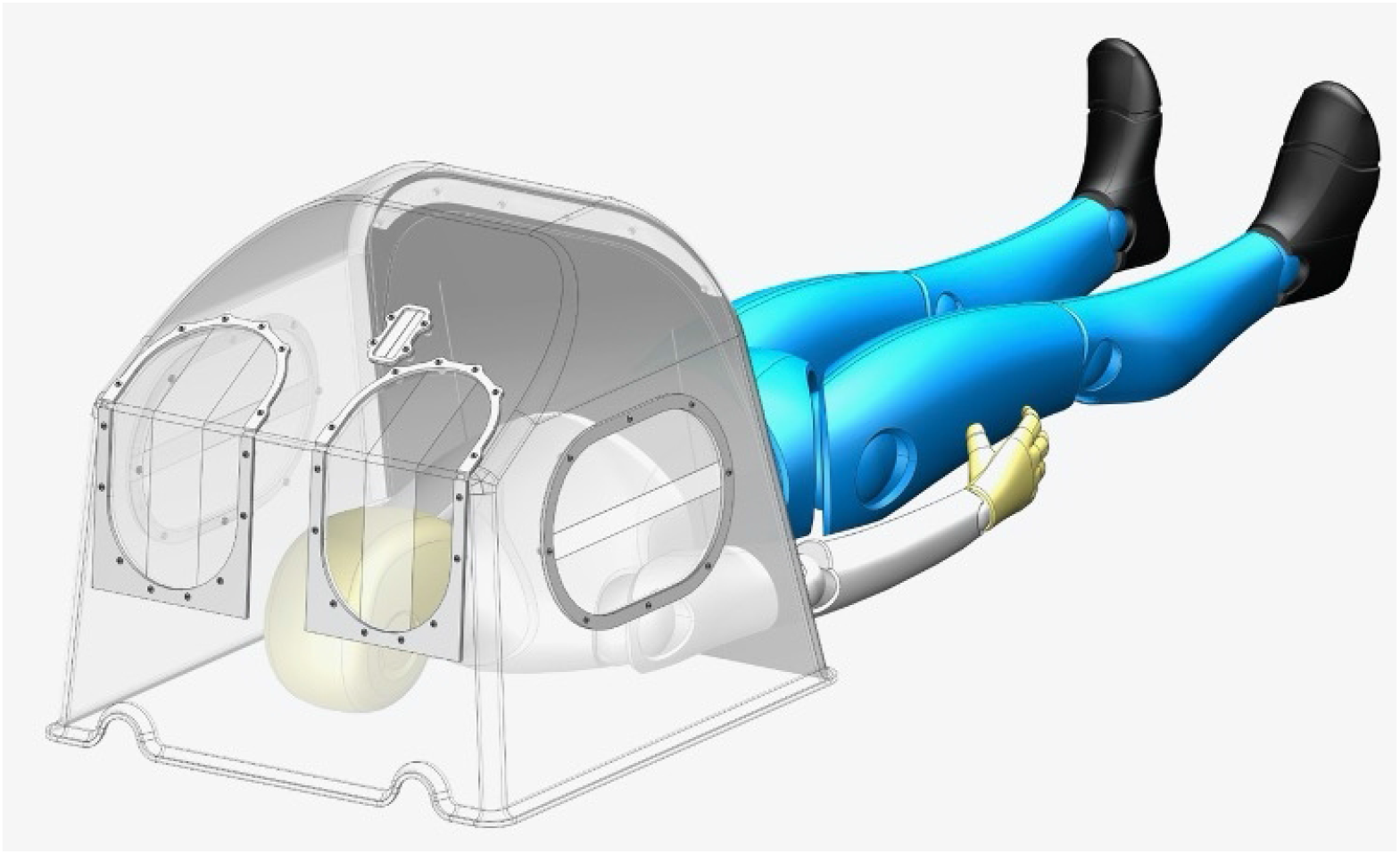
Polyethylene terephthalate glycol-modified (PETG) vacuum formed shield with overlapping silicone access ports. The overlaps for the operator are vertical and hortizontal for the assistant to maximise operators arm freedom and movement.

### Computational Fluid Dynamics

We used ANSYS Fluent software^6^ to perform Computational Fluid Dynamics modelling of aerosol and droplet dispersion. A total of 1000 particles ranging in size from 1 to 500 microns, were modelled during an initial cough followed by a normal breath^7^. In addition to the standard Navier-Stokes equations governing the three-dimensional features of the fluid (conservation of mass, momentum and energy), the simulation also uses a Discrete Phase Model that tracks the individual particles in Lagrangian coordinates^8^. The interaction of the particles with the airflow is modelled as a one-way coupling and applied as a post-processing exercise. This means the flow affects the momentum and energy of the particles, but the surrounding fluid flow remains unaffected by the motion of particles.

Particle paths are determined by their size. Heavier particles have more momentum and therefore behave under projectile motion, traveling until they are trapped by a surface. Lighter particles are affected by velocity streamlines, circulating until either encountering a surface or being extracted. Parameters used by the model include ambient temperature, breath temperature, cough cone angle detailed in the supplemental material.

### Experimental modelling

We modelled multiple scenarios to ascertain the optimal design of the shield. We then modelled the dispersion of particles emitted from a cough with the shield and without the shield in a standard sized side room.

#### Experiment 1: Suction tube position

To determine the effect and optimal position of suction within the box, we placed a suction tube modelled on a typical Yankauer sucker either on the patient’s chest or vertically aligned and in-line with the patient’s head. The model was run in each of these position at a suction flow rate of 40 L min^−1^. Figure 2 shows particle size, streamline velocity and position of trapped particles (see supplementary material for videos).

**Figure 2.**
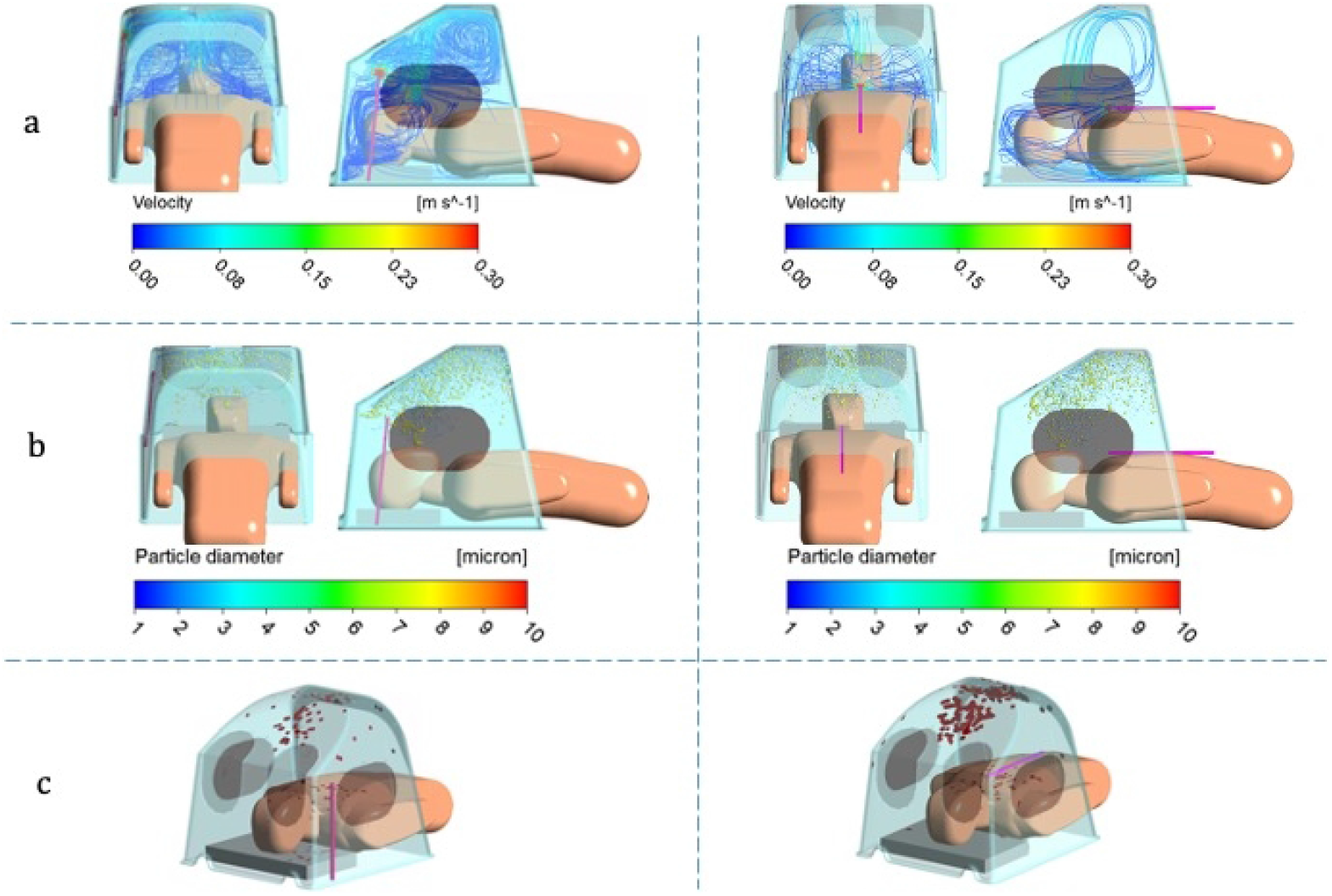
Computational Fluid Dynamics model with suction placed either vertically and in-line with patient’s head (left) or on chest (right) (a) Airborne cough particle tracker (b) velocity streamlines from pressure outlet (c) trapped particles on internal shield surfaces

#### Experiment 2: Effect of additional openings

The shield is designed to allow ambient air to be drawn in around gaps in the curtain which lies over the torso/upper abdomen of the patient, allowing entrainment of air and extraction of box environment through the suction. If the shield is used to perform procedures, further air is likely to enterand there is potential for aerosol to escapevia gaps created by the operator’s arms (figure 3). We modelled the shield without additional openings and then a worst-case scenario analysis with all openings patent.

**Figure 3.**
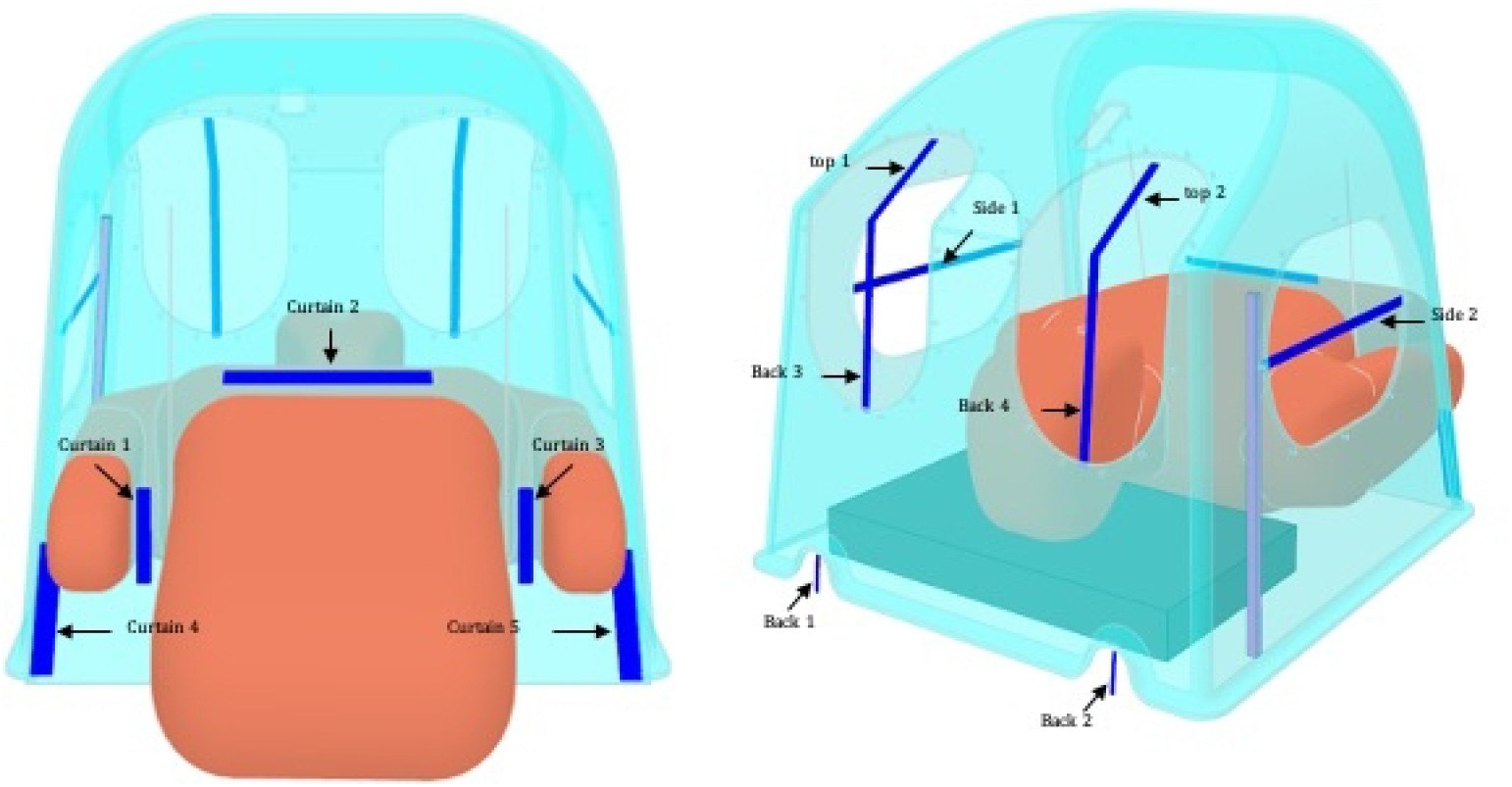
Location of potential openings in shield: Back 1 & 2 are ports for ventilation tubing and suction. All other potential openings are covered with silicone flaps. Air enters if these flaps are opened for access to patient.

#### Experiment 3: Suction flow rate

Suction flow rate depends on the amount of negative pressure produced by the vacuum source, the resistance of the suction system, and the viscosity of the substance (in this case, air). We modelled a range of suction values (40–160L min^−1^) to assess the impact of suction rate on the performance of the shield. We also modelled the effect of these suction ranges with and without additional openings as detailed in Experiment 2.

#### Experiment 4: Presence vs absence of shield

We modelled presence of the shield at the lowest suction rate (40L min^−1^) with the maximum number of openings patent (worst case scenario) vs no shield, in a room measuring 4m x 4.5m x 3.1m with an air change rate of 10ach^9^ and an ambient temperature of 23°C. Cough particle velocity of 5ms^−1^ and cough cone angle of 30 degrees^10,11^. Supply and exhaust grills were positioned as per pictures in the supplementary material. We modelled this scenario with three staff positioned around the patient.

### Results

#### Experiment 1: Suction tube position

The position of the Yankeaur suction tip within the shield altered the air flow. With the suction placed on the patient’s chest, after 500 seconds, 31% particles were trapped on the shield wall, 56% particles were extracted through the suction tube, 7% remained floating within the box while 6% escaped. Compared to the suction catheter placed vertically, in line with patient’s head, fewer particles were extracted and more particles were trapped on the patient’s body and shield walls as the air was drawn towards the patient’s chest. With the suction placed in line with the patient’s head, 23% more particles were extracted through the suction tube, with ∼1% remained floating within the shield (Figure 4).

**Figure 4.**
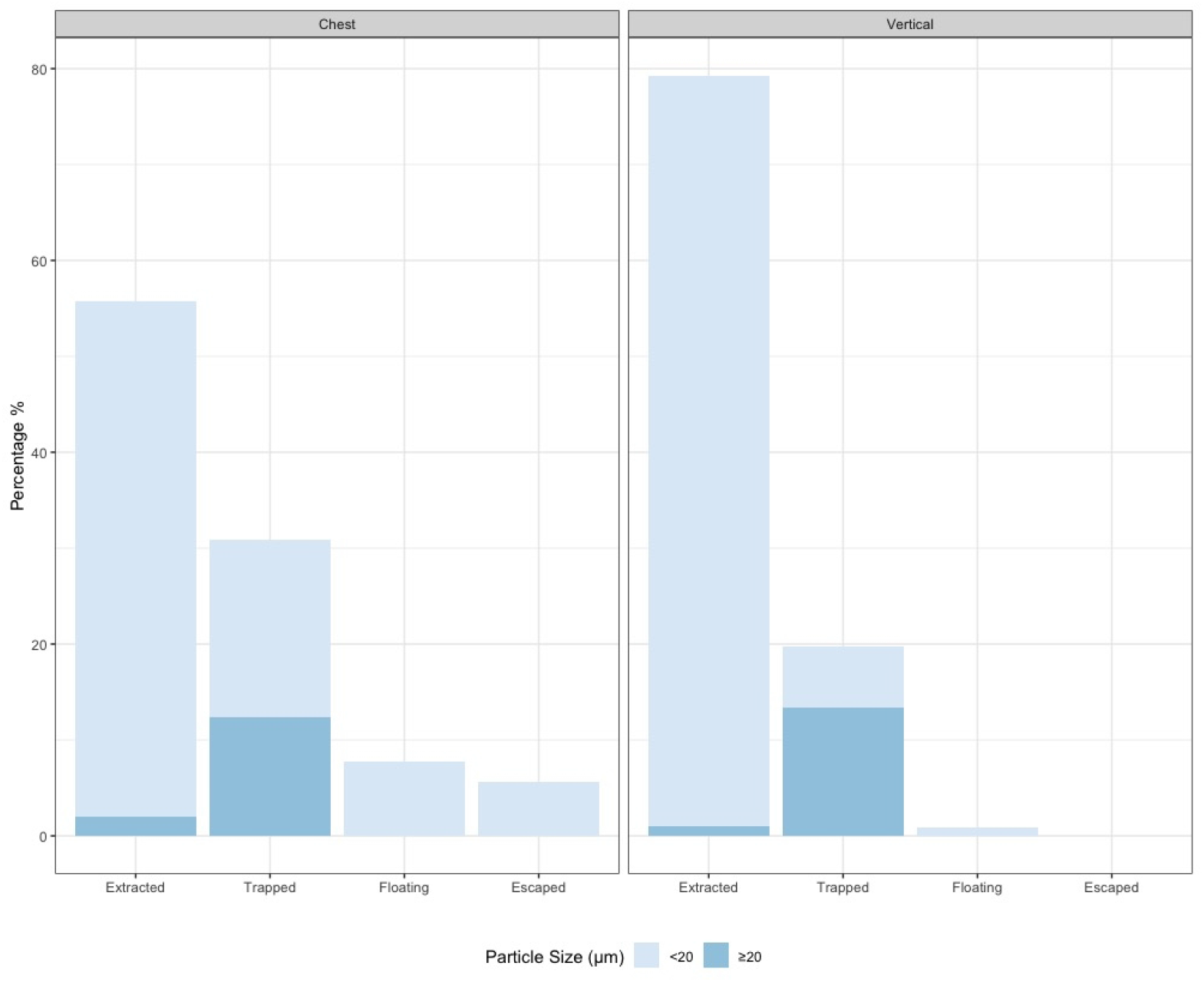
End location of particles after 500 seconds, according to size and suction position either on paitent’s chest (Chest) or vertically, in-line with their head (Vertical). Extracted = Extracted through suction tube, Trapped = trapped on internal surface of shield Floating = remaining inside shield in aerosol, Escaped = escaped perimeter of shield.

#### Experiment 2: Suction flow rate

With the shield placed on the patient, the number of particles escaping the box was negligible over a range of suction flow rates from 40–160 L min^−1^ with 99% of particles being removed via suction or trapped on the walls of the shield. (Figure 5 left panel)

**Figure 5.**
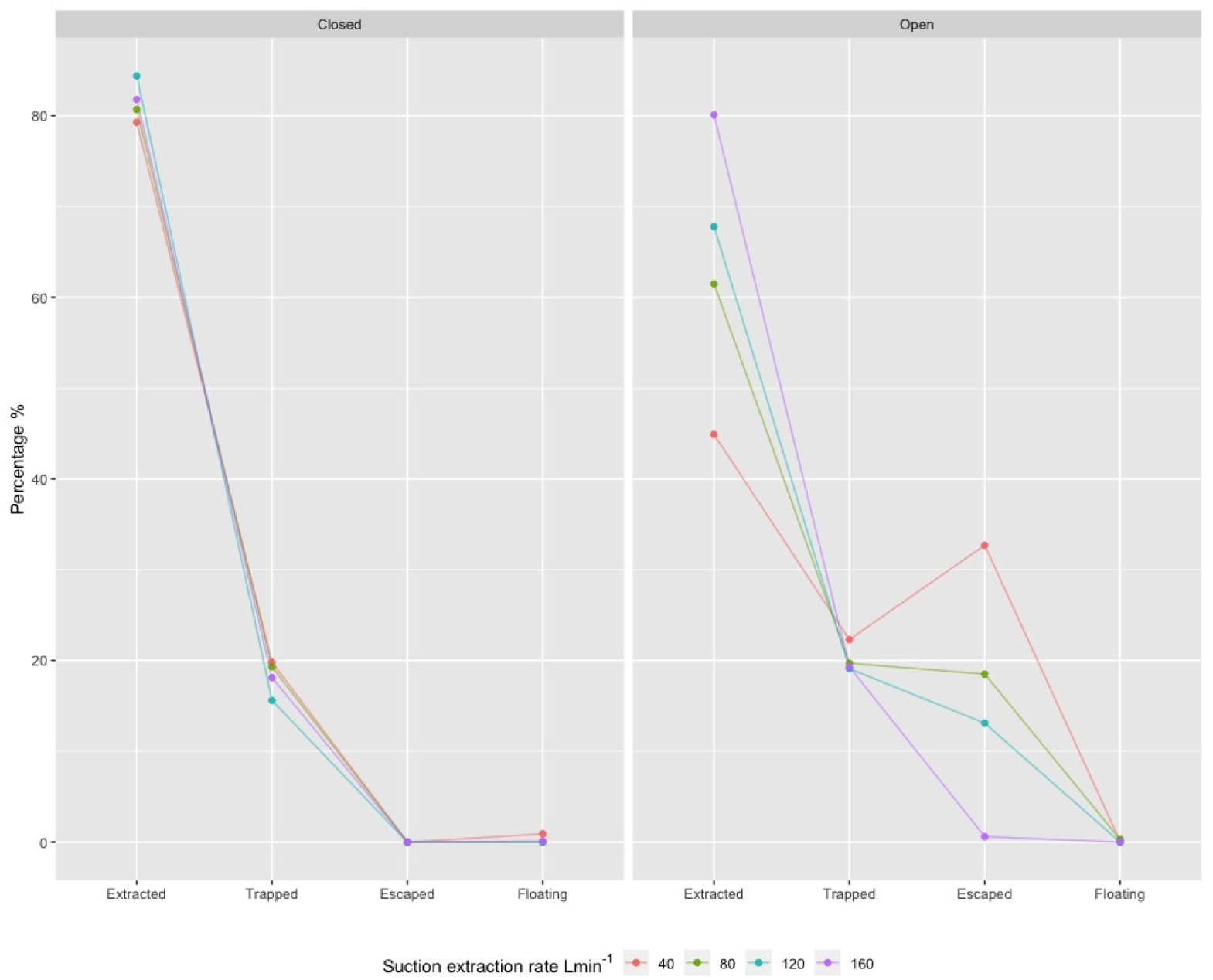
End locations of particles after 500 seconds in closed model (left panel) and open model – all potential opening fully patent (right panel) according to suction rate.

#### Experiment 3: Impact of incomplete seals

With all openings patent, particle escape from the shield ranged from 1 – 33% depending on the suction flow rate (figure 5 right panel). The proportion of particles trapped on the walls is less affected by incomplete seals (16–20% in closed model, 19–22% in open model). The proportions of particles by size, escaping from each opening, in the open and closed model, at varying suction rates can be found in supplementary material.

#### Experiment 4: Presence versus absence of shield

Without the shield, larger particles travelled along their trajectory until they landed either on the medical staff or surfaces in the room. Smaller particles travelled along the paths created by the velocity streamlines from the room ventilation system, these particles circulated in the room until they were either trapped on surfaces inside the room or extracted via room ventilation.

Figure 6 shows the positions of particles, by size, using room parameters outlined in the methods. The shield performance is improved inside the positively pressurised room, keeping more particles contained within due to the air flow pattern around the shield and the increased pressure gradient between the room and the shield.

**Figure 6.**
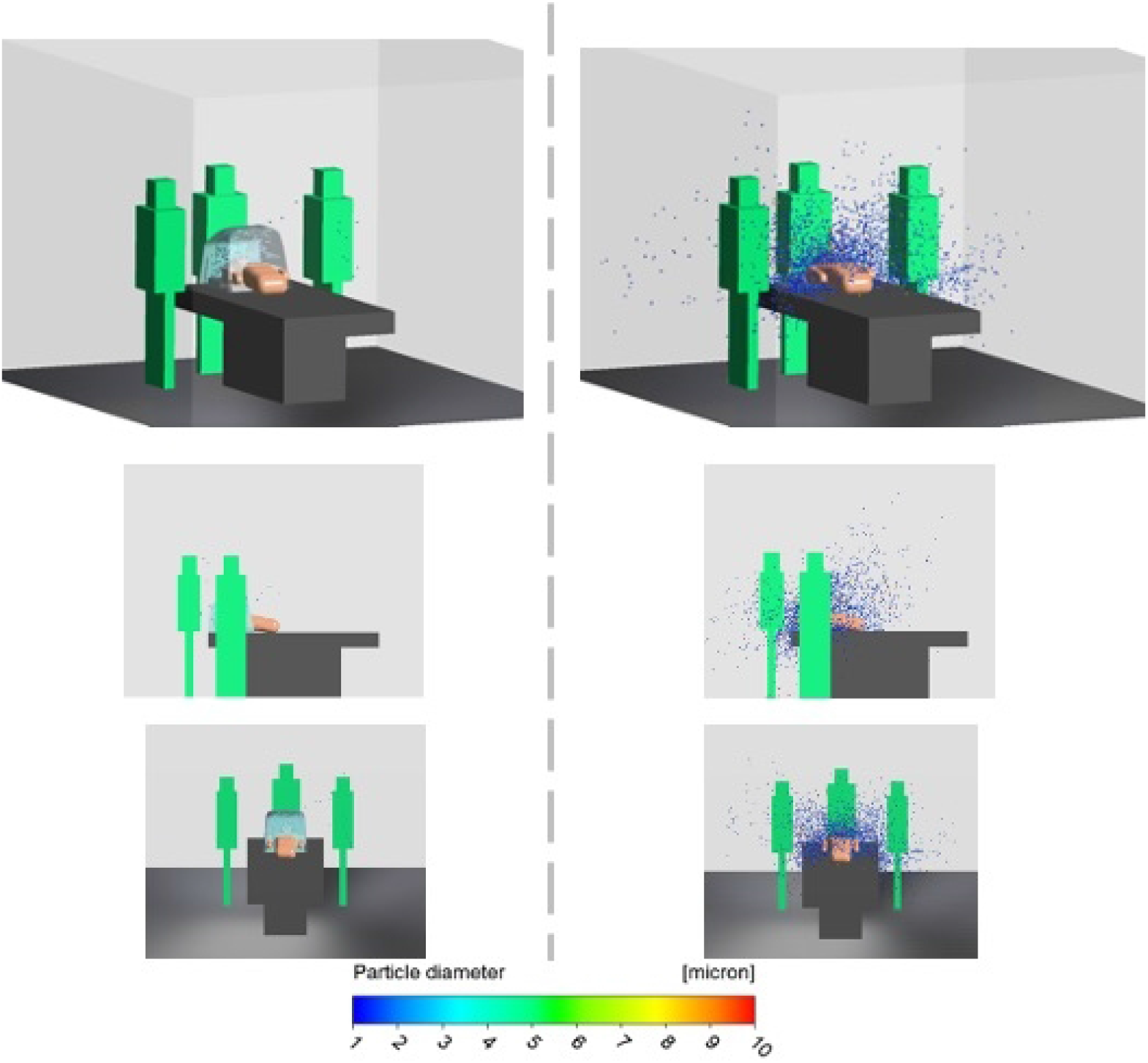
Distribution of particles by size in room with 10ach following a cough then normal breathing. Left panel: modelled with shield on, all potential openings patent and suction inside shield of 40 L min^−1^. Right panel: same room without shield present.

### Discussion

This is the first study to use CFD to evaluate the effect of a barrier shield on aerosol and droplet trajectories. The CFD study shows that this shield is highly effective at removing both droplets and aerosolised particles which would otherwise contaminate the local environment and personnel.

The Covid-19 pandemic has highlighted the risk of transmission via aersolisation and respiratory secretions. HCWs are at highest risk of contracting infections transmitted in aerosols and droplets when performing Aerosol Generating Procedures (AGPs) and working in high risk clinical areas including intensive care, operating theatres, endoscopy and bronchoscopy units^12^. Up to 4.4% patients with Covid-19 in China were health care workers or individuals who worked in medical facilities^13^. The figure is higher in Italy (8%) and Spain (11.6%)^14, 15^. Compliance with PPE is advised to reduce transmission rates^16^; however, despite complying with Public Health England guidance, healthcare workers remain vulnerable to droplet contamination^17^.

There is growing interest in, and use of, barrier devices as a method to reduce occupational exposure and potentially nosocomial infections during these types of procedures. Similar barrier methods (termed as ‘intubation protection box’) have been used to compare direct laryngoscopy, videolaryngoscopy, and video-laryngoscopy using a protective intubation box, in an airway mannequin in vivo^18^. In this simulated intubation scenario, a mucosal atomisation device was used to simulate a cough and aerosolisation of droplets, which was attached to a 10ml syringe containing a red-dye solution. While direct and videolaryngoscopy were associated with dye being deposited on the laryngoscopist’s faceshield, gown, arms, glove, neck, and hair, use of the box reduced the deposition of dye only on the gloves and forearms within the box. No dye was visible on any part of the laryngoscopist located outside the box. While this approach primarily quantifies the spread of droplets, it is unlikely to characterise the distribution of fine aerosols.

Our study adds quantitative data to support the plausibility of such barriers reducing environmental and personal biocontamination. We suggest such barriers could be used as an extra precaution in addition to gold standard PPE as part of a PPE bundle to reduce nosocomial and occupational infections. These findings reinforce the pressing need for the systematic assessment of new devices that may offer definitive protection before their clinical deployment. We used state of the art computational fluid dynamics modelling, which has a robust track record in complex industrial design that has frequently superseded the need for in vivo confirmation. This approach afforded the modelling of a range of scenarios that cannot be replicated in vivo. Modelling the distribution and locations of particles in a room beyond the shield depends on factors including but not limited to; room size, ventilation location, rate and turbulence and the position of equipment and personnel within the room. While these factors vary widely between hospitals and even within hospitals, the exact proportions and final locations of particles will naturally vary. Nonetheless, our study clearly demonstrates the utility of this device in containing infectious matter regardless of the environment in which it is used.

While our study is limited by not assessing the effect of this shield in vivo, we modelled conservatively. We modelled all potential opening fully patent reducing the efficacy of the shield. The seal formed by the silicone flaps around the operators arms reduces potential openings. Furthermore, it is unlikely that all openings modelled will be patent, for example the airway assistant will either be accessing patient from right or left rather than both sides. Additionally, when used in a positively pressurised room the performance is further improved. The results presented in the open model are therefore a worst-case scenario. A further limitation is that we did not consider the evaporation of the particles, however this is likely to decrease the time particles reside within the shield. Although the extraction of the majority of particles occurred through the suction, use without suction will trap still trap 97% of droplets produced.

As with all new equipment, training and familiarity is vital, and all staff involved should be confident in both the theory and practical application. In summary, we provide robust computational fluid dynamics modelling data showing that this custom design shield effectively minimises exposure of healthcare workers to droplets and aerosols that may transmit hazardous pathogens, and that environmental contamination is reduced. Further clinical assessment is warranted, as the targeted use of such devices may enhance the PPE armamentarium to healthcare workers globally in preparation for future predicted pandemics.

## Data Availability

All data is available online.

http://www.the-mtc.org/news-items/intubation-shield-supporting-our-frontline-nhs-workers

## Authors’ contributions

Drafting Manuscript: PP

Data Analysis: PP, AP

CFD Model and Figures: MT, EH, AP

Revision and approval of final version of article: all authors

## Deceleration of Interest

Designers/Patent holders: PP/IR

## Acknowledgments

Rolls Royce and The Manufacturing Technology Center were instrumental in the rapid design, development, production and distribution of the shield.

Andy York, Head of Programmes, Manufacturing Technology, Rolls Royce.

Alan Pardoe Partnership Manager – The Manufacturing Technology Centre, Rolls-Royce Danny McGee, Chief Engineer, The Manufacturing Technology Centre (MTC), Coventry. Multimatic – a global enterprise supplying engineered components, systems and services to the automotive industry, who developed the vacuum forming tooling for the shield.

Aston Martin – for help with distribution of the shields for testing.

Edith Blennerhassett Director, Arup – an independent firm of designers, planners, engineers, architects, consultants and technical specialists who supported this study gratis.

## Appendices (if applicable)

**Figure 7.**
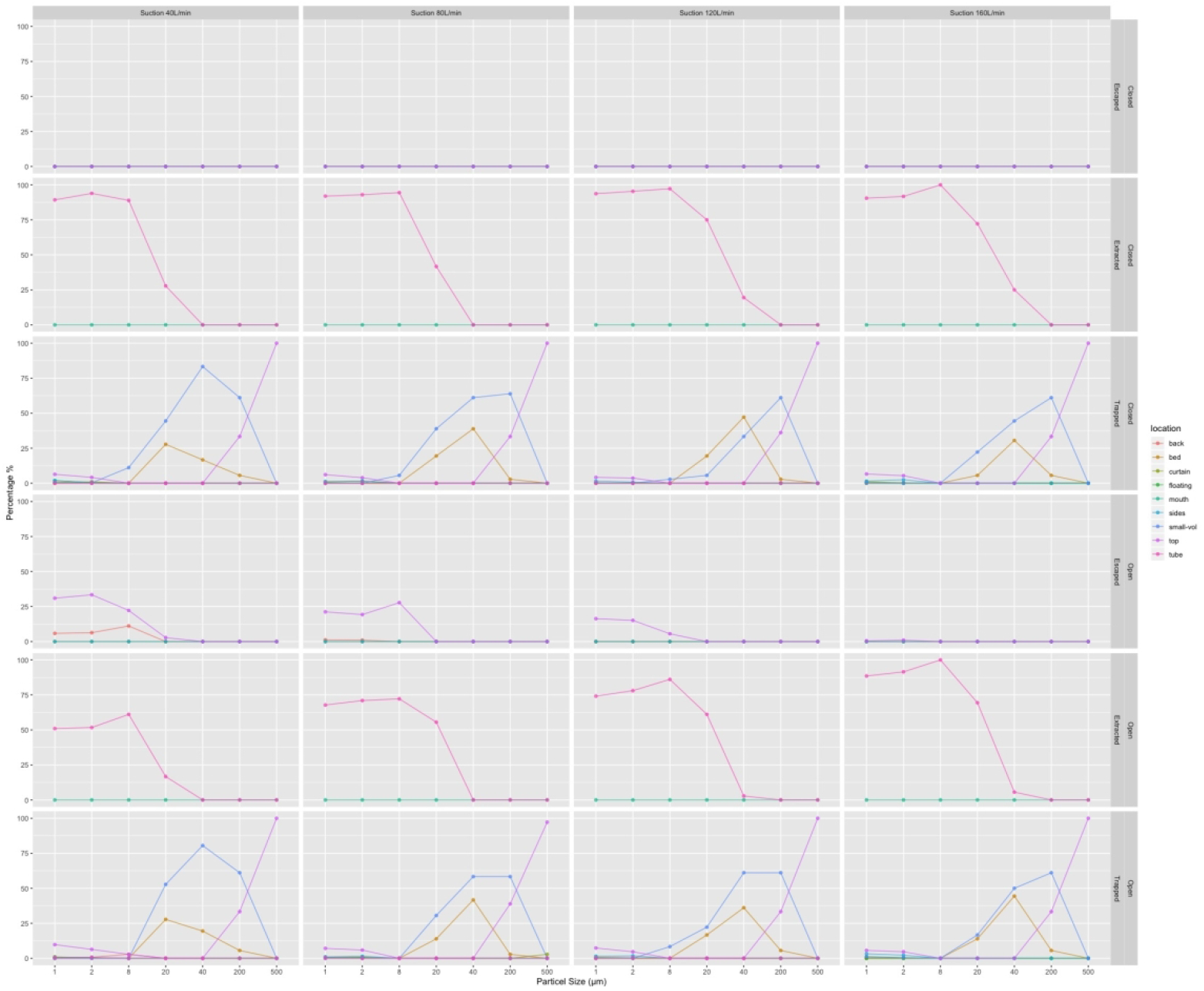
Locations of particles by size after 500 seconds. Faceted by suction rate, model (open vs close) and fate of particles.). Extracted = Extracted through suction tube, Trapped = trapped on internal surface of shield Floating = remaining inside shield in aerosol, Escaped = escaped perimeter of shield. For locations of openings see figure 3.

